# Integrative analysis of cfDNA features from ultra-low coverage whole genome sequencing enables robust detection of ovarian cancer

**DOI:** 10.64898/2026.07.02.26357095

**Authors:** Z. Hanzlíková, J. Styk, O. Pös, O. Biró, S. Bokorová, L. Lukyová, J. Sitarčík, T. Sládeček, W. Krampl, A. Mészáros, P. Hunyadi, Sz. Máté, Á. Égető, J. Rigó, T. Sedláčková, J. Radvánszky, J. Budiš, T. Szemes

## Abstract

**Background:** Despite advances in circulating tumor DNA analysis, reliable detection of oncological disease from ultra-low coverage whole genome sequencing (ulcWGS) remains challenging, particularly at low tumor fractions. This study leverages cell-free DNA (cfDNA) characteristics to develop and evaluate a robust, integrative binary predictive model for ovarian cancer (OC) status screening. OC represents a growing global burden and is often diagnosed at advanced stages due to the lack of specific early symptoms and effective screening strategies, highlighting the need for sensitive and broadly applicable early detection approaches.

**Methods and Findings:** We analyzed plasma cfDNA from OC patients (*N* = 85) and cancer-free controls (*N* = 41) using ulcWGS (∼1×). Participation in the study was voluntary, and all participants provided written informed consent before any study-related procedures under study approval No. 16119-8/2022/EÜIG. Within an integrated workflow combining standardized laboratory processing, bioinformatic pipelines, and machine learning (ML), we extracted 21 features capturing copy number variations (CNVs) and fragmentomic characteristics to identify complementary signatures distinguishing OC from controls. Predictive models were developed using XGBoost with hyperparameter optimization and evaluated on an independent test set (*n* = 25% of the cohort). A dual-threshold classification strategy was applied to define an uncertainty zone and optimize screening performance.

CNV-derived and fragmentomic features assessed in exploratory analysis on the training-validation set showed moderate discriminative power (AUC 0.569 – 0.946) but substantial overlap between groups. On the test set, the CNV-only model achieved an AUC of 0.855 (sensitivity 85%, specificity 50%), while the fragmentomics-only model reached an AUC of 0.8825 (sensitivity 95%, specificity 30%). Both feature domains captured complementary aspects of tumor-derived cfDNA, with fragmentomics favoring sensitivity and CNV-derived metrics improving specificity. Integration of both feature classes improved performance, yielding an AUC of 0.900, sensitivity of 85.00%, and specificity of 90.00%. SHAP analysis confirmed contributions from both feature types without a single dominant predictor.

**Conclusions:** We present an integrative cfDNA framework for OC detection based on ulcWGS that combines CNV and fragmentomic signals to improve diagnostic performance over single-feature approaches. By enabling robust detection of tumor-associated patterns at ultra-low sequencing depth, this approach demonstrates that meaningful cancer discrimination can be achieved without reliance on deep sequencing. This highlights the potential of cost-effective and scalable liquid biopsy strategies for population-level cancer screening their integration into personalized and preventive oncology.

## Introduction

Ovarian cancer (OC) remains the most lethal gynecological malignancy, affecting women of all ages, predominantly those over 50 [1, 2]. OC encompasses a heterogeneous group of subtypes that differ in their molecular features, clinical behavior, and therapeutic response. Approximately 90% of cases are epithelial in origin, with major subtypes including high-grade serous ovarian cancer (HGSOC), low-grade serous ovarian cancer (LGSOC), clear cell carcinoma, mucinous carcinoma [3], and endometrioid ovarian cancer (EOvC) [4]. In addition to epithelial tumors, less common histological types include malignant germ cell tumors, which occur more frequently in younger women and may exhibit aggressive clinical behavior [5], as well as sex-cord stromal tumors [6].

Among these, HGSOC is the predominant subclass, constituting nearly 70% of OC cases [7] and is associated with the highest lethality. It is an aggressive form of epithelial OC, frequently characterized by a clinically silent and insidious onset that makes early detection challenging. The absence of noticeable symptoms in the initial phases of disease development, coupled with the lack of practical screening and diagnostic tests, contributes to delayed diagnosis in most OC patients. While the 5-year survival rate can reach up to ∼90% when confined to the ovaries (Stage I), and remains around 70% when limited to the pelvis (Stage II), this rate drops sharply to 20% or less once the disease has spread beyond the pelvis (III -IV) [8]. Despite this survival gradient, only a minority of OC cases are diagnosed at early stages (I – II), with approximately 66% of patients presenting with advanced disease at diagnosis [9, 10]. Conventional serum biomarkers, such as CA-125, lack the sensitivity and specificity required for screening asymptomatic women, being undetectable in ∼50% of early-stage cases. In addition, CA-125 levels are frequently elevated in benign conditions and non-ovarian malignancies and undetectable or modest levels in some OC subtypes [11, 12]. The rapid growth and invasive spread of HGSOC often lead to diagnosis at developed stages, significantly contributing to its unfavorable prognosis [13]. Consequently, minimally invasive liquid biopsy approaches have gained substantial attention for early cancer screening, as they enable repeated sampling with minimal burden [14]. In this context, genome-wide interrogation of cell-free DNA (cfDNA) captures tumor-associated alterations and offers the potential to detect signals from multiple malignancies within a single assay [15].

Among the available strategies, whole genome sequencing (WGS) of plasma cfDNA provides a broad perspective to detect tumor-derived signals without prior knowledge of mutational hotspots. While ultra-low coverage WGS (ulcWGS), typically operating around ∼0.1 – 1× coverage, does not offer the resolution of deep sequencing [16], its genome-wide scope enables the integration of multiple complementary signals, such as copy number variations (CNVs) [17, 18], which represent a well-established component with broad diagnostic and clinical applications [19], and fragmentomic features of cfDNA [20]. Fragmentomics broadly encompasses the genome-wide analysis of cfDNA fragmentation patterns, capturing features such as fragment length profiles and their genomic distribution, where deviations reflect tumor-associated chromatin alterations. This is particularly relevant for ovarian tumors, which frequently harbor extensive large-scale copy number gains and losses [21–23]. cfDNA fragmentation patterns of OC can highlight these changes through the detection of anomalies in size and distribution of cfDNA fragments. Tumor-derived cfDNA reflects the disordered chromatin structure and dysregulated nuclease activity of malignant cells, resulting in patterns that differ from the regular, nucleosome-guided profiles seen in healthy plasma. Importantly, only a small fraction of plasma cfDNA is tumor-derived, while the majority originates from normal tissues, diluting tumor-specific signals [24]. In particular, cancers are frequently associated with a reduced median fragment length (insert size) and an increased proportion of short and ultra-short fragments [25]. Such alterations in fragment size dynamics form an essential component of the signal leveraged by cfDNA-based analyses [26]. Both genome-wide fragmentomics and CNVs capture distinct but complementary aspects of tumour-derived alterations and their relation to health and cancer conditions [27]. However, detecting and interpreting early-stage cancers using low coverage sequencing data remains analytically challenging [28]. Sparse and irregular read distributions, coverage fluctuations [29], GC content biases [30, 31], technical variation in insert size [32, 33], and a range of sequencing-run artefacts together with other batch-specific, non-biological sources of variability [34], can reduce detection of true tumor-associated signals from the technical noise. Rather than explicitly modeling or correcting these sources of bias, we employed an integrative approach designed to capture robust patterns that persist despite such variability These sources of bias were not explicitly modeled or corrected in this study. Instead, we employed an integrative modeling approach intended to learn robust patterns in the data, potentially reducing the influence of such confounding effects without directly parameterizing them. Reliable interpretation of pathological and physiological cfDNA signatures thus requires sophisticated bioinformatic pipelines, standardized analytical workflows, and integrative modeling to mitigate these constraints [35].

Recognizing that single-biomarker solutions lack sufficient robustness for early cancer screening [36, 37], complex information may be frequently missed in asymptomatic populations. Recent work has thus shifted toward multi-feature frameworks testing in parallel for distinct cfDNA variability. Fragmentomics, in particular, offers a potentially widespread screening paradigm [38] that makes it easier to implement and more acceptable for the general populations [28]. However, its utility is maximized when integrated with genome-wide profiling and additional metrics.

We hypothesized that integrating genome-wide structural imbalance with fragmentation dynamics would capture complementary aspects of tumor-derived cfDNA biology detectable at ultra-low sequencing depth. By building on established tools and concepts for CNV profiling, fragmentomic ctDNA burden estimation, fragmentation features, and sequencing-level quality control, we derive in-house features that are subsequently integrated using machine learning (ML) modeling to differentiate OC patients from healthy controls. Such a multi-feature approach overcomes the limited sensitivity of standalone biomarkers and yields more stable predictive performance across heterogeneous clinical presentations [39]. We demonstrate strong predictive performance of the combined model on the independent test set (AUC = 0.989), exceeding that of individual biomarkers, which showed moderate discriminative ability (AUC 0.569 – 0.946) in exploratory analyses on the training-validation set. These results highlight the benefit of integrating diverse cfDNA-derived features and support the potential of this approach for preventive cancer screening.

## Methods and materials

### Ovarian cancer prospective cohort

The study enrolled *N* = 41 healthy controls without known ovarian pathology and *N* = 85 OC patients. Written informed consent was obtained from all participants. Samples were collected under approval number 16119-8/2022/EÜIG between 27 July 2022 and 22 October 2025 at the First Department of Obstetrics and Gynaecology, Semmelweis University, Budapest, Hungary (Fig. 1). Healthy controls were excluded if they had a known germline BRCA1/2 mutation, a family history of ovarian or breast cancer, gynecological pathology or an ongoing pregnancy. Among the oncological cohort, *N* = 82 patients provided a baseline pre-surgery blood sample collected in the presence of the primary tumor, whereas three patients contributed only a single post-surgery follow-up sample without a corresponding baseline specimen. In total, *N* = 33 follow-up samples were obtained, of which *n* = 30 were paired with matched baseline samples from the same individuals, enabling limited longitudinal analyses. Patient classification was based on established clinical and pathological criteria. OC cases were defined by histopathologically confirmed OC following surgical treatment and staged according to the FIGO classification. Classification was determined by the treating gynecologic oncology team using routine clinical, surgical and histopathological findings prior to molecular analysis. The cancer-free group comprised healthy women matched to the patient cohort based on key demographic characteristics, including age. At baseline collection, the mean age of cancer patients was 60.43 years compared with 56.30 years in controls. According to the FIGO (International Federation of Gynecology and Obstetrics) staging system, the entire oncological cohort (*N* = 85) included Stage I (*n* = 5; 5.9%), Stage II (*n* = 9; 10.6%), Stage III (*n* = 51; 60%), and Stage IV (*n* = 9; 10.6%) cases, with 11 patients (12.9%) lacking stage information (S1 Fig.).

**Fig 1.**
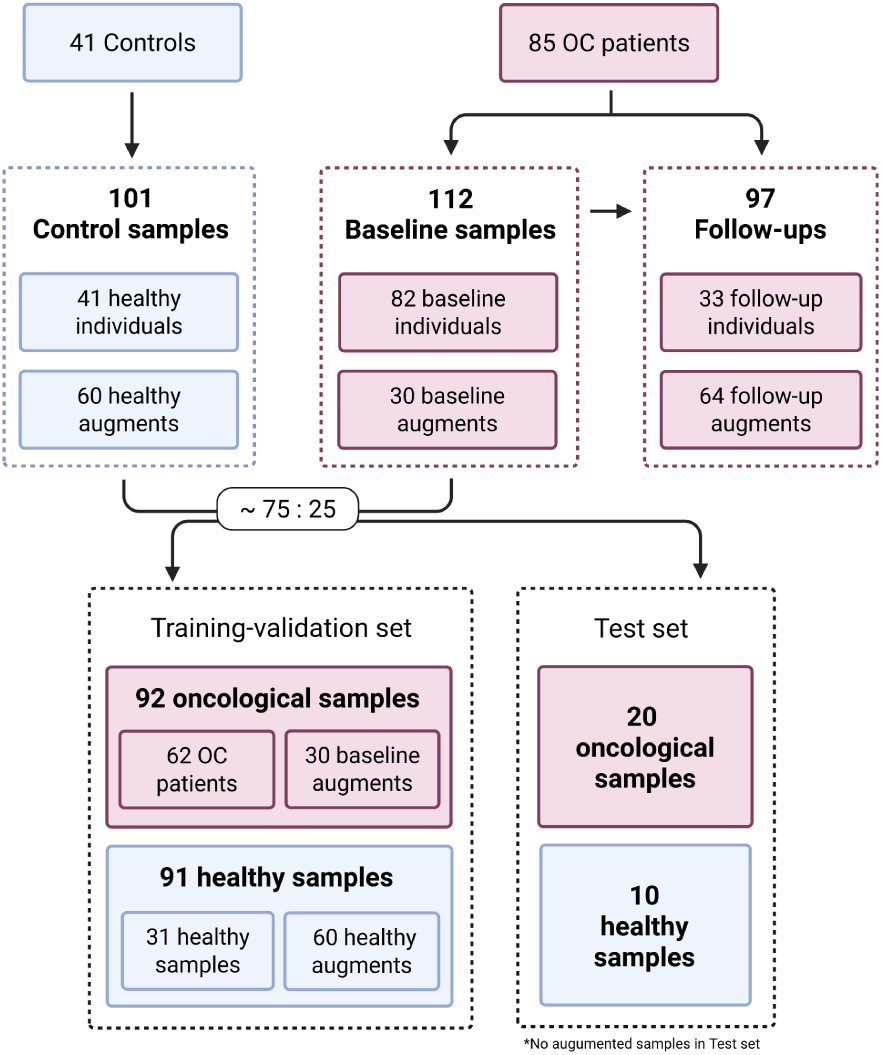
Overview of cohort composition, sample augmentation, and dataset splitting.

### Sample collection, processing and sequencing

Prospectively collected venous blood samples, ∼9 ml of each, were taken into Vacuette® K2EDTA tube (Greiner Bio-One GmbH) at the First Department of Obstetrics and Gynaecology, Semmelweis University (Budapest, Hungary) and kept at 4°C until processing. Samples were anonymized and transferred to the laboratory of Clinomics Europe Ltd. (Budapest, Hungary). Blood samples were centrifuged to plasma within 4 hours of collection by a 2-step protocol at 1,150g for 10 min (4°C), followed by 12,500g for 10 min (4°C). To ensure sample quality, plasma samples were screened for contamination by free hemoglobin using spectrophotometric quantification on a NanoDrop One Spectrophotometer (Thermo Fisher Scientific). Samples with oxygenated hemoglobin levels >0.8 mg/ml (absorbance peak at 414 nm wavelength) were classified as hemolyzed and were excluded from downstream analyses [40]. All of the sequenced samples had an oxyhemoglobin level below 0.3 mg/ml. Plasma aliquots were cryopreserved at −80°C and shipped to the Genomics Core Facility at the Comenius University Science Park (CU SP; Bratislava, Slovakia) under controlled, temperature-stable conditions on dry ice. To prevent sample mix-ups at any stage of the process, each sample was registered in the electronic system upon its arrival at the CU SP laboratory.

Total cfDNA extraction was performed using 680 µl of clarified plasma with the QIAamp DNA Blood Mini Kit (QIAGEN) following the manufacturer’s instructions and the concentration of cfDNA in the plasma was determined using the Qubit dsDNA HS Assay Kit (Thermo Fisher Scientific) according to the manufacturer’s guidelines. Then, 30 μl of isolated cfDNA per sample was subjected to WGS library construction using the TruSeq® Nano DNA Library Prep kits (Illumina) and TruSeq DNA CD indexes (Illumina). Subsequently, libraries were amplified for 8−9 PCR cycles, pooled equimolarly to 1,000 pM, and sequenced in a short-read 100 bp paired-end setup on the NextSeq 2000 system (Illumina) under an ulcWGS workflow targeting ∼1× coverage, with observed average depth ranging from 0.99 to 2.85× (S1 Table). Given the prospective study design and gradual sample collection, samples were received and processed in five batches containing cancer and cancer-free samples. To mitigate technical variability and batch effect, all ulcWGS data were generated using uniform laboratory protocols, identical reagent kits, and the same sequencing platform.

In addition, for a subset of samples, sufficient biological material was available to generate multiple aliquots, which were processed and sequenced independently. These technical replicates provided naturally augmented versions of the same underlying specimen (Fig. 1). Rather than being treated merely as replicates, these augments were explicitly leveraged in downstream analyses as additional training instances. This strategy allowed us to partially compensate for the imbalance between the oncological and control cohorts while preserving biological consistency. Importantly, because the augments originate from the same source material but undergo independent library preparation and sequencing, they capture realistic technical variability without introducing artificial noise. As such, they contribute to the increased robustness of statistical and ML models and improve generalization.

### Bioinformatics processing

Sequenced reads underwent trimming with Cudapt 4.4 [41] to retain only informative fragments as follows. Quality trimming cut-off was set to 20 for both ends and both pairs, and then reads longer than 500 had their excess bases removed. Subsequently, we discarded: reads whose expected number of errors exceeded 3, reads with more than 10 N bases, and reads shorter than 35 bases. The filtered reads were then aligned to the GRCh38 reference genome using BWA-MEM 0.7.17 [42]. Following alignment, PCR duplicates were marked with Picard 3.0.0 [43] to minimize amplification bias and ensure the reliability of downstream analyses. A total of 21 cfDNA-derived features characterizing CNV and fragmentomic profiles were then extracted using tools and methods described below (Table 1).

**Table 1.** Summary of cfDNA ulcWGS-derived features used in the study.

| Tool | Feature | Description |
| --- | --- | --- |
| WisecondorX [44, 45] | cnv_count | Total number of CNV events. |
|  | cnv_sum_length | Cumulative genomic span affected by CNVs. |
|  | cnv_sum_level | Sum of absolute CNV levels across all detected CNV segments, reflecting overall copy number deviation magnitude. |
|  | cnv_sum_ratio | Sum of absolute normalized read-depth ratios across all CNV segments, capturing aggregate copy number signal intensity. |
|  | cnv_sum_len_level | Length-weighted sum of absolute CNV levels, integrating both CNV magnitude and affected genomic span. |
|  | cnv_sum_len_ratio | Length-weighted sum of absolute normalized read-depth ratios across CNV segments. |
|  | cnv_sum_len_level_ratio | Composite metric integrating CNV length, level, and normalized ratio to represent overall genomic instability. |
|  | CPA_score | Copy number profile abnormality score [45]. |
|  | std_ratios_per_chrom | Standard deviation of ratios per chromosome. |
|  | med_seg_variance | Median segment variance per bin [44]. |
| Fraggle [46] | ctDNA_Burden | Composite fragmentomic score summarizing global deviations in cfDNA fragment length distributions relative to healthy reference profiles, reflecting the overall burden of tumor-associated fragmentation [46]. |
| Qualimap [47] | insert_size_score_delfi | Genome-wide insert-size asymmetry score capturing global fragmentation irregularities via ratio of short (100 – 150 bp) and long (151 – 220 bp) reads [48]. |
|  | median_insert_size | Median insert size of paired-end cfDNA fragments mapped to autosomes. |
|  | mean_insert_size | Mean insert size of paired-end cfDNA fragments mapped to autosomes. |
|  | std_insert_size | Standard deviation of cfDNA insert sizes across autosomes. |
|  | ultra_short_reads_prop | Proportion of cfDNA fragments <100 bp mapped to autosomes. |
|  | short_read_prop | Proportion of cfDNA fragments within the range of 100 – 150 bp mapped to autosomes. |
| | shorter_read_prop | Proportion of cfDNA fragments $\leq 150$ bp mapped to autosomes. |
|  | long_reads_prop | Proportion of cfDNA fragments within the range of 151 – 220 bp mapped to autosomes. |
|  | ultra_long_reads_prop | Proportion of cfDNA fragments >220 bp mapped to autosomes. |
|  | longer_read_prop | Proportion of cfDNA fragments >150 bp mapped to autosomes. |

### Copy Number Variations

CNVs were called using WisecondorX [44] with a bin size of 20 kbp, accounting for marked duplicate reads, while all other parameters were kept at their default settings. CNV-derived features were analyzed at the genome-wide scale using the complete set of events without additional filtering, jointly considering both copy number gains and losses to capture overall structural genomic instability. As WisecondorX requires a healthy reference cohort for CNV prediction, a reference set of 20 healthy individuals was randomly selected from the dataset, ensuring a balanced representation across all sequencing run batches. Each individual was included only once in the reference, even when augmented samples were available.

#### Fragmentomics

Fragmentomic profiles were characterized using a combination of fragment length-based and read-based metrics. Fragle [46], a deep learning-based tool, was applied to genome-wide cfDNA fragment length distributions to estimate the proportion of ctDNA and capture cancer-associated fragmentation patterns. Complementary insert size-based metrics were obtained using Qualimap 2.2.2d [47], with all features calculated exclusively from autosomal reads only to avoid bias from mitochondrial DNA and ensuring consistency across samples. Together, the Fragle and Qualimap derived metrics provide a comprehensive representation of cfDNA fragmentation, integrating both tumor-informed signatures and general fragmentomic characteristics to enable detailed analysis of structural properties of circulating DNA.

### Predictive model development and evaluation

#### Sample selection

Due to the unpredictable behavior of post-surgery follow-up samples, which is closely associated with different sample stages, varying time intervals from surgery to collection (ranging from 0 to 351 days, mean 100,3 days), different chemotherapy management and the absence of multiple consecutive sampling points, assessing their status regarding the presence and extent of oncological disease is challenging. This also prevented the formation of sufficiently large and homogeneous groups for reliable trend analysis. Therefore, for the OC predictive model development, we considered only pre-surgery oncological samples and cancer-free individuals. The post-surgery samples were evaluated separately.

All OC subtypes were evaluated jointly in this analysis. Despite their distinct mutational landscapes, we aimed to identify global cfDNA fragmentomic signatures and overall mutational burden metrics that capture shared biological characteristics across histological subtypes rather than subtype-specific alterations.

#### Data augmentation and dataset splitting

Individuals were divided into a training-validation set and an independent test set in an approximately 75:25 ratio relative to the original cohort (*N* = 82 oncological and *N* = 41 healthy individuals), while maintaining a balanced age distribution. The training-validation set comprised *n* = 62 oncological and *n* = 31 cancer-free individuals, whereas the test set included *n* = 20 oncological and *n* = 10 cancer-free individuals. Sample counts reflect the inclusion of augmented samples (*n* = 92 oncological vs. 91 control) in the training-validation set, while the independent test set consisted exclusively of original, non-augmented samples to ensure unbiased evaluation, with each individual contributing only a single measurement (Fig 1). Age was explicitly balanced between oncological and control groups and preserved across the training-validation and test sets to minimize potential age-related confounding. Without such balancing, the model could misinterpret age-associated biological variation as disease-related, leading to increased false-positive predictions among older healthy individuals, or alternatively, fail to detect disease if its expression differs in older populations. Furthermore, because age is often correlated with diagnosis in oncology, an imbalanced design could lead the model to rely on age as a surrogate predictor (“higher age = higher risk”) rather than learning disease-specific biological patterns.

To further prevent result bias, ensured that none of the individuals included in the test set were used as part of the WisecondorX reference for feature derivation.

The study includes ovarian cancer patients and healthy controls with baseline and follow-up samples. Augmented samples were used exclusively in the training-validation set, while the independent test set contained only original samples. Created in https://BioRender.com

#### Machine-learning modeling and validation

We applied the Leave-One-Group-Out (LOGO) cross-validation strategy on a training-validation data set, a specific form of cross-validation where one group of samples is used for validation while all other groups are used for training. In this case, a group consisted of all samples from the same individual, ranging from 1 to 8 depending on the number of augmented samples.

For the predictive model, we selected XGBoost due to its strong performance on structured, heterogeneous data and its ability to provide interpretable feature importance scores [49]. Model hyperparameters that influence classification performance were systematically optimized during the training process using a Bayesian optimization framework implemented in Optuna [50], specifically employing the Tree-structured Parzen Estimator (TPE) sampler. A total of 100 optimization trials were performed, during which key hyperparameters including the number of estimators, learning rate, maximum tree depth, subsampling ratio, column sampling ratio, gamma, minimum child weight, and L1/L2 regularization terms, were sampled from predefined ranges. Within each fold, imbalance between classes was handled adaptively by recalculating class weights according to the label proportions in the training data. For each modeling iteration, predictions were evaluated using the default decision threshold of 0.5. After completing the training and validation process, a single optimal decision threshold was subsequently determined on the entire training-validation subset, with an emphasis on achieving at least 90.00% sensitivity, reflecting a conservative strategy. In regions where predicted probabilities for healthy and oncological classes overlapped, this approach prioritized capturing as many oncological cases as possible, even at the cost of flagging a larger number of healthy samples for preventive follow-up. Final performance metrics on the training-validation set were computed as the average performance across all validation groups.

Inspired by practical usage in clinical screening centers, where specific biological or technical factors may lead to indecisive results, we also adopted a dual-threshold approach to classification. In addition to the primary decision threshold optimized for high sensitivity, we implemented an alternative secondary threshold to achieve a minimum specificity of 80.00%. This allowed us to define an “uncertain zone”, a range of predicted probabilities between the primary and secondary thresholds, representing predictions with increased uncertainty. All downstream analyses of predictive performance were conducted both including and excluding samples falling within this grey zone. For transparency, we also report the coverage parameter, defined as the proportion of samples assigned a definitive prediction outside the uncertain zone, providing an estimate of the effective applicability of the model.

#### Feature evaluation

To better understand the distinct contributions of different cfDNA-derived signals, we first trained separate predictive models for CNV-derived features and fragmentomic metrics. This approach allowed us to evaluate the independent discriminative power of structural and fragmentomic information in isolation. Subsequently, all features were combined into a single integrated model to examine their joint predictive performance and potential synergistic effects within this comprehensive framework. Feature contributions were interpreted using SHAP (SHapley Additive exPlanations) values, which provided a quantitative measure of each metric’s influence on the model predictions, highlighting how CNV and fragmentomic signals collectively drive classification performance [51].

## Results

### Evaluation of CNV-derived features

We first evaluated the discriminative potential of CNV-derived features independently (Fig. 2). CNV-derived metrics demonstrated systematic shifts between OC and control samples, reflected by moderate to high AUC values ranging from 0.623 to 0.946. These shifts are consistent with increased genomic instability and tumor-associated copy number alterations (CNA) contributing to the circulating cfDNA pool. However, despite differences in central tendency, substantial distributional overlap persisted between groups, indicating that individual CNV metrics alone do not provide complete discrimination at the single-feature level. A subset of *n* = 20 healthy samples was used to train the WisecondorX internal reference. Since these samples exhibited CNV profiles and AUC values comparable to the rest of the healthy cohort (S2 Fig.), we included the full set of healthy samples in our analysis to maximize statistical power.

**Fig 2.**
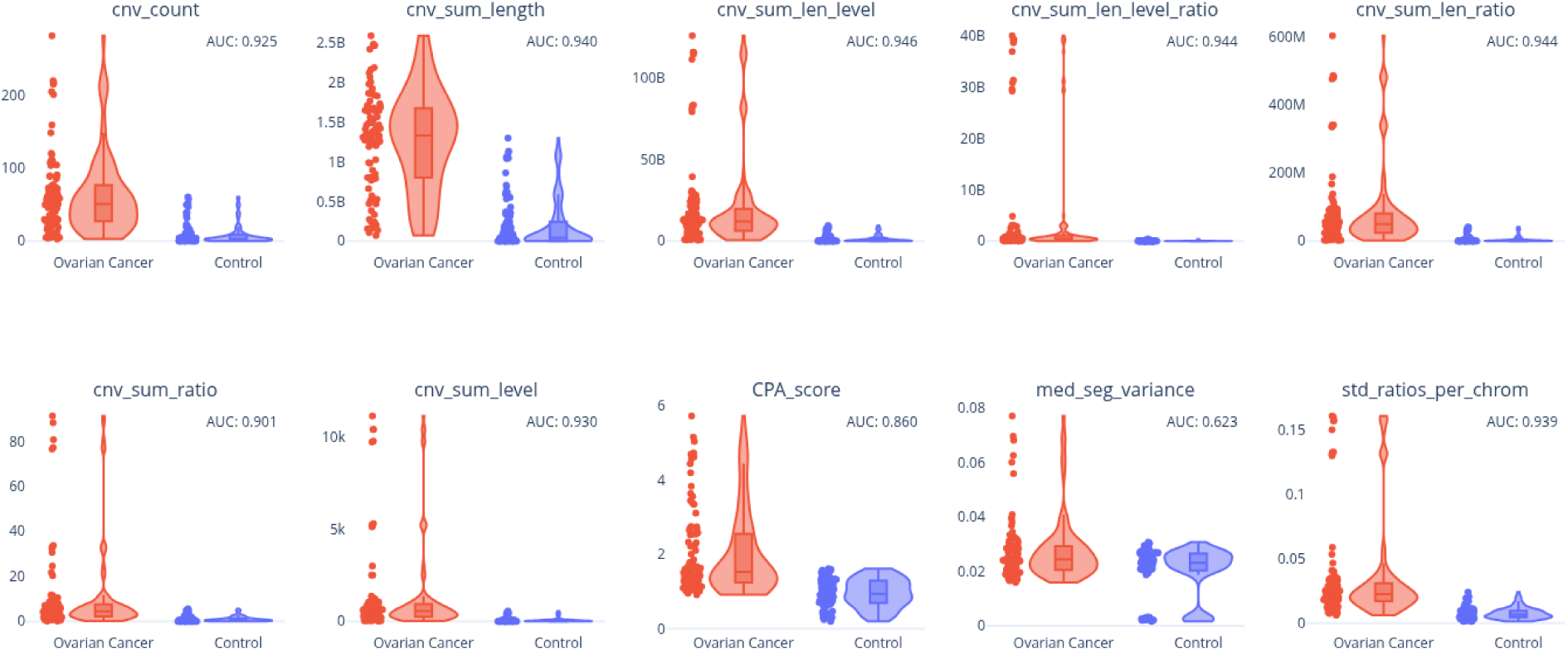
Differences of CNV-derived metrics between cancer-free controls (Control, including reference samples) and ovarian cancer samples. Each violin represents the full distribution of values within the respective group.

To further assess the multivariate structure of CNV-derived features, principal component analysis (PCA) was performed (Fig. 3). The PCA scatter plot revealed partial separation between OC and control samples, primarily along PC1, suggesting that CNV-associated alterations contribute substantially to the dominant variance structure. Nevertheless, a considerable proportion of samples remained overlapped, indicating that not all variance is disease-driven and that linear combinations of CNV metrics alone are insufficient to achieve clear diagnostic separation.

**Fig 3.**
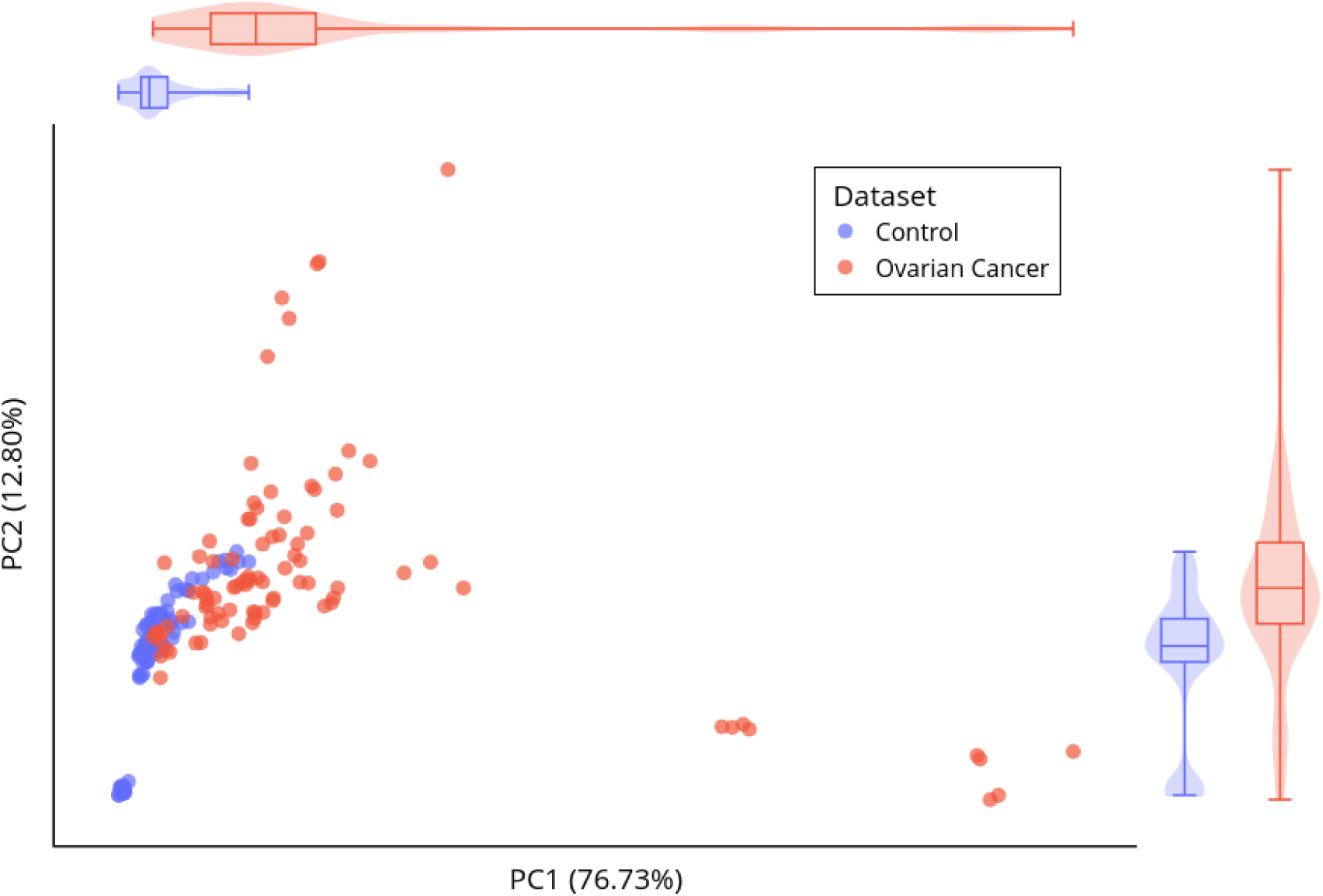
PCA projection of all CNV-derived metrics. The first two principal components (PC1 and PC2) together explained 89.53% of the overall variability in the CNV feature space.

We subsequently trained a supervised ML model using the full set of CNV-derived features. On an independent test set, the CNV-only model achieved an AUC of 0.855, accuracy of 73.33%, sensitivity of 85.00%, and specificity of 50.00% without applying a zone of uncertainty. When introducing an uncertainty zone threshold, 3.33% of samples were excluded, while accuracy and sensitivity improved modestly to 75.86% and 89.47%, respectively, and specificity remained unchanged at 50.00%.

### Evaluation of fragmentomics features

We next evaluated fragmentomic features independently using the same analytical framework. Fragmentomic metrics exhibited measurable group-specific differences, consistent with altered cfDNA fragmentation patterns in cancer patients. After aligning metric directionality, features achieved discriminative performance with AUC values ranging from 0.569 to 0.871 (Fig. 4), indicating that tumor-derived cfDNA contributes detectable structural signatures. However, similar to the CNV-derived metrics, substantial overlap between healthy and cancer samples was observed, highlighting that individual fragmentomic parameters capture only part of the disease-associated signal.

**Fig 4.**
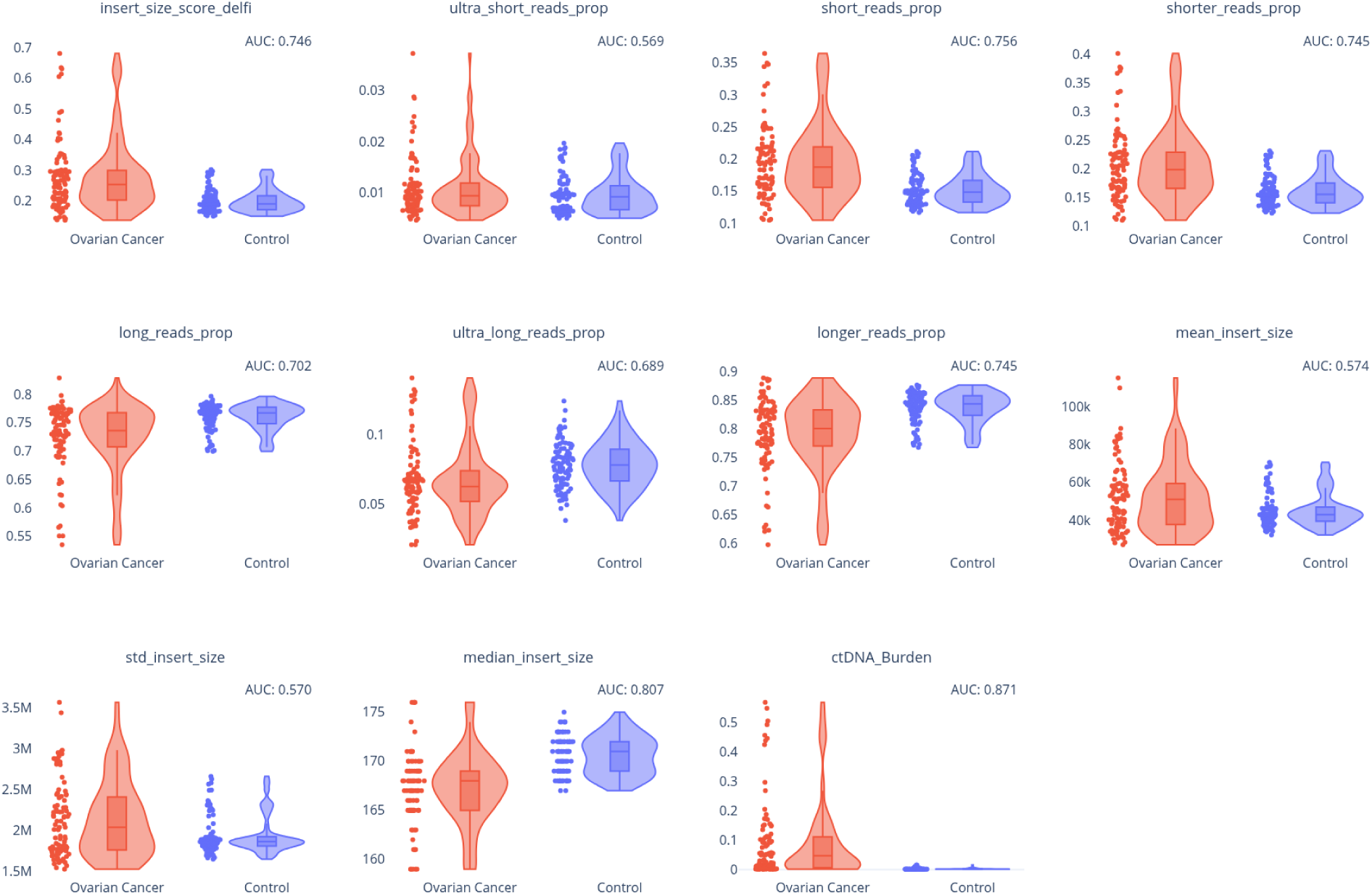
Violin plots of fragmentomic metrics comparing healthy controls (including reference samples) and ovarian cancer samples. Each violin represents the distribution of values within the respective group, with boxplots and individual data points shown. The AUC for each metric is reported to quantify its discriminatory capacity.

To evaluate the global variance structure, PCA was performed on all fragmentomic features (Fig. 5). The PCA projection demonstrated partial separation between OC and control samples, indicating that fragmentation patterns contribute to disease-associated variance. Nonetheless, extensive overlap between groups persisted, suggesting that the dominant axes of variance include both biological heterogeneity and shared background variability.

**Fig 5.**
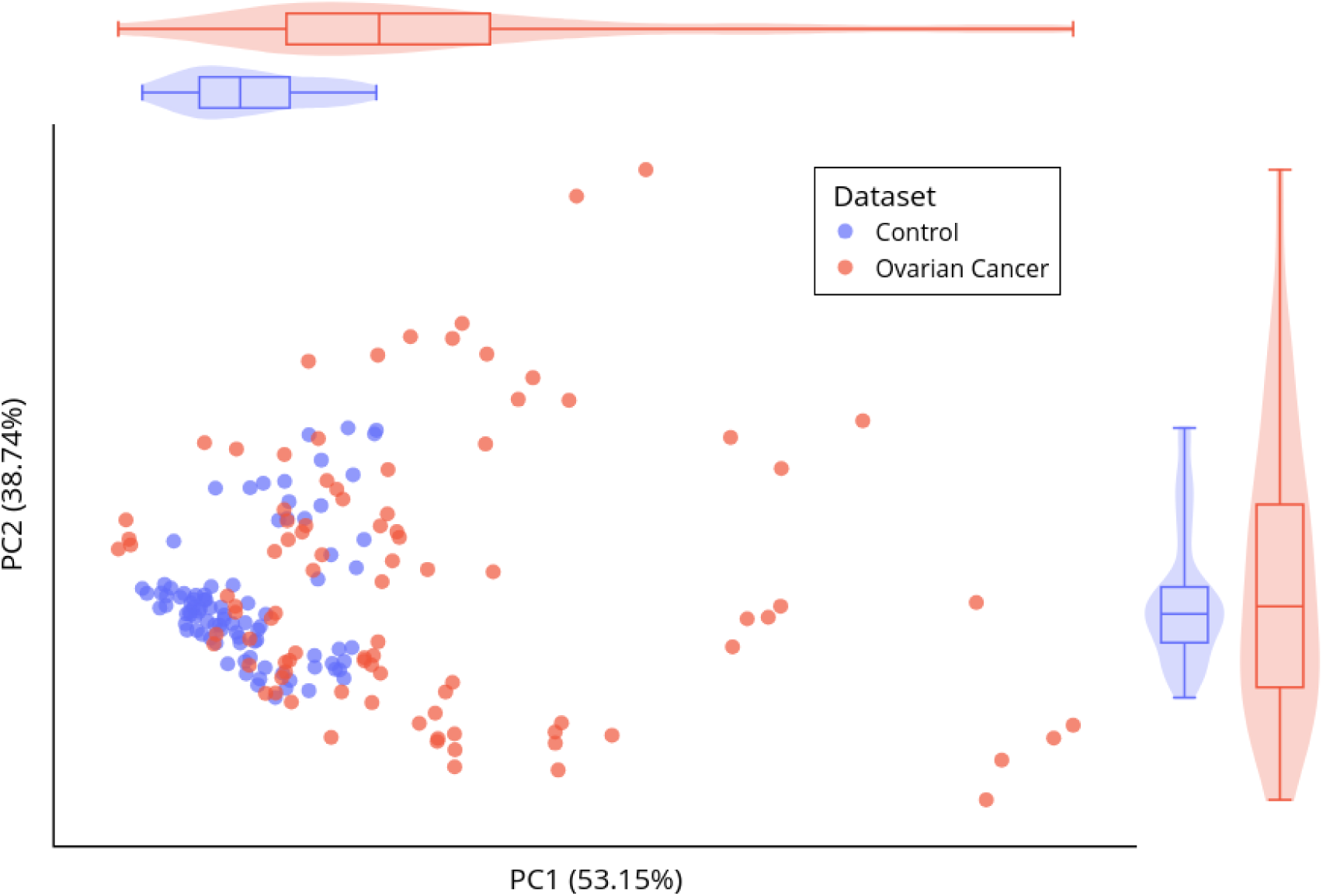
PCA of fragmentomic metrics. The first two principal components (PC1 and PC2) together explain 91.89% of the total variance in the fragmentomic feature space.

A supervised ML model trained exclusively on fragmentomic features improved discriminative performance compared to individual metrics. On an independent test set, the fragmentomic-only model achieved an AUC of 0.8825, accuracy of 73.33%, sensitivity of 95.00%, and specificity of 30.00% without applying a zone of uncertainty. When introducing an uncertainty zone threshold, 13.33% of samples were excluded, while accuracy and specificity improved substantially to 80.77% and 42.86%, respectively, and sensitivity slightly decreased to 94.74%.

### Combined evaluation of CNV-derived and fragmentomics features

Taken together, both CNV-derived and fragmentomic feature sets contain biologically meaningful signals associated with OC. At the univariate and PCA levels, both domains demonstrate partial group separation but substantial overlap, reflecting biological heterogeneity and shared variance components. Supervised ML models enhance classification performance by integrating multiple complementary features within each domain, translating subtle molecular differences into clinically relevant discrimination. These results indicate that while the CNV-only model and fragmentomics-only model detect most oncologic cases, their ability to correctly classify non-oncologic samples is limited.

After evaluating CNV-derived and fragmentomic metrics independently, we next investigated their combined behavior within a unified feature space to determine whether integration of both biological domains improves discrimination and reveals additional structural relationships. By merging CNV and fragmentomic features, we aimed to assess whether OC-associated cfDNA alterations manifest as coordinated multidimensional shifts rather than isolated changes within a single feature class.

We first examined the global variance structure using PCA applied to the full set of cfDNA-derived features (Fig. 6), together capturing 77.34% of overall variability. Compared to PCA performed separately on CNV or fragmentomic features, the combined projection demonstrated slightly improved group-level separation between OC and control samples. The primary axis of variation reflected a mixture of CNV-related amplitude shifts and coordinated fragmentomic alterations, suggesting that both feature classes contribute to shared disease-associated variance. Nevertheless, partial overlap between groups persisted, indicating that OC-associated signal is distributed across multiple orthogonal components and cannot be fully resolved by linear dimensionality reduction alone.

**Fig 6.**
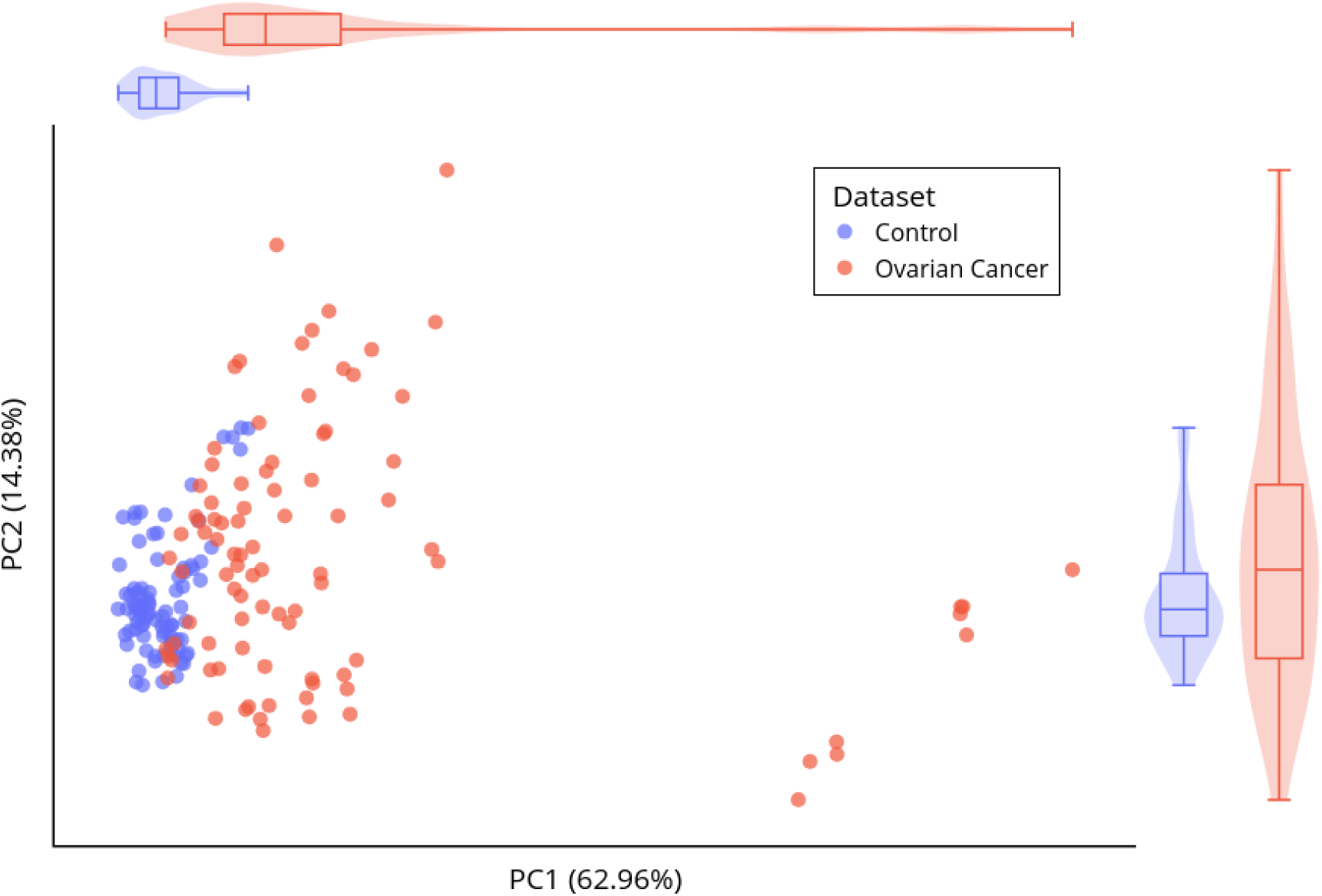
PCA of all metrics, including both CNV- and fragmentomics-derived features. The first two principal components (PC1 and PC2) together explain 77.34% of the total variance.

To further investigate the intrinsic structure of the integrated feature space prior to supervised modeling, hierarchical clustering was performed using two complementary distance metrics on data that were transformed to account for compositional structure (for proportional features) and then standardized to ensure comparable feature scales across all variables: Euclidean distance with Ward’s linkage (Fig. 7A) and correlation-based distance (1 − Pearson correlation) with complete linkage (Fig. 7B). Continuous non-proportional features were standardized using z-score transformation to ensure comparable scaling and to prevent dominance of features with larger numerical ranges. Despite standardization, the two distance metrics preserve fundamentally different geometric interpretations. Euclidean distance captures absolute differences in feature magnitude and remains sensitive to global shifts in cfDNA signal intensity, whereas correlation-based distance emphasizes similarity in feature profiles independent of absolute scale, highlighting coordinated structural patterns across CNV and fragmentomic metrics.

**Fig 7.**
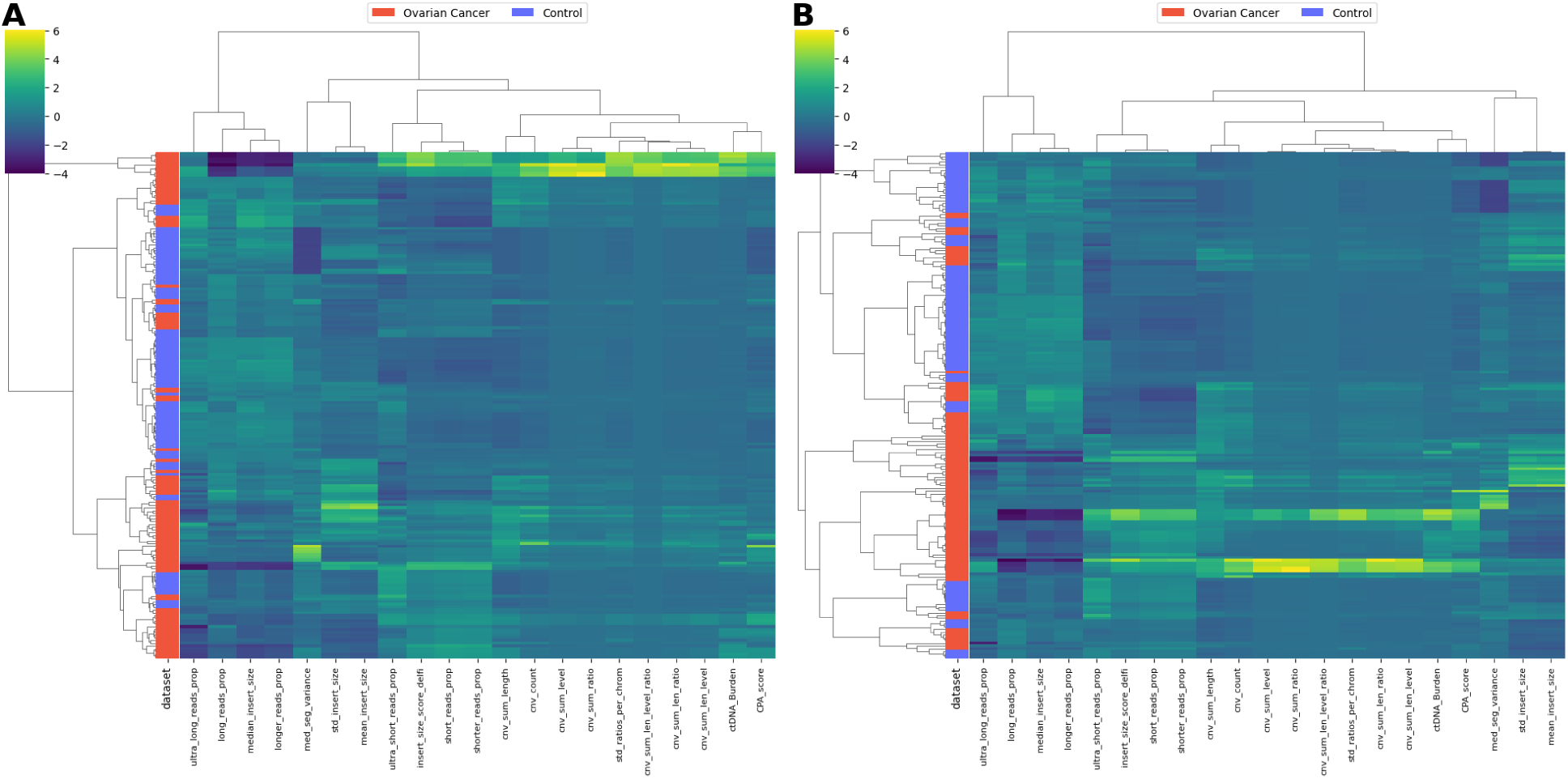
Clustering of cfDNA features using complementary distance metrics. Hierarchical clustering of samples based on cfDNA-derived features. Heatmaps were generated using (A) Euclidean distance with Ward’s linkage and (B) correlation-based distance (1 − Pearson correlation) with complete linkage.

Both clustering approaches revealed partially overlapping yet non-identical cluster structures. Euclidean-based clustering emphasized magnitude-driven separation, with subsets of OC samples characterized by globally elevated CNV instability and altered fragmentation parameters. In contrast, correlation-based clustering identified groups of samples sharing similar multivariate feature profiles even when overall signal intensity differed. Examination of the feature-feature correlation matrix demonstrated non-trivial interdependencies between several CNV and fragmentomic metrics, revealing modular structures within the combined feature space. Distinct feature clusters emerged, some enriched for CNV amplitude-related metrics and others dominated by fragmentation-pattern features, indicating partial redundancy within domains but complementary organization across domains. These findings suggest that sample separation is not driven solely by global amplitude differences but rather by complex, multidimensional relationships among features.

Given this structured yet overlapping multivariate landscape, we subsequently trained a supervised ML model using the full set of CNV and fragmentomic features simultaneously (Fig. 8). By allowing nonlinear integration of complementary signals, the model can capture higher-order interactions and conditional dependencies not apparent in univariate analyses or linear projections. On an independent test set, the combined-feature model without applying the zone of uncertainty achieved an AUC of 0.9000, accuracy of 86.67%, sensitivity of 85.00%, and specificity of 90.00%. The combined model effectively integrates complementary information from the individual CNV and fragmentomic models, leveraging their respective strengths in sensitivity and specificity.

**Fig 8.**
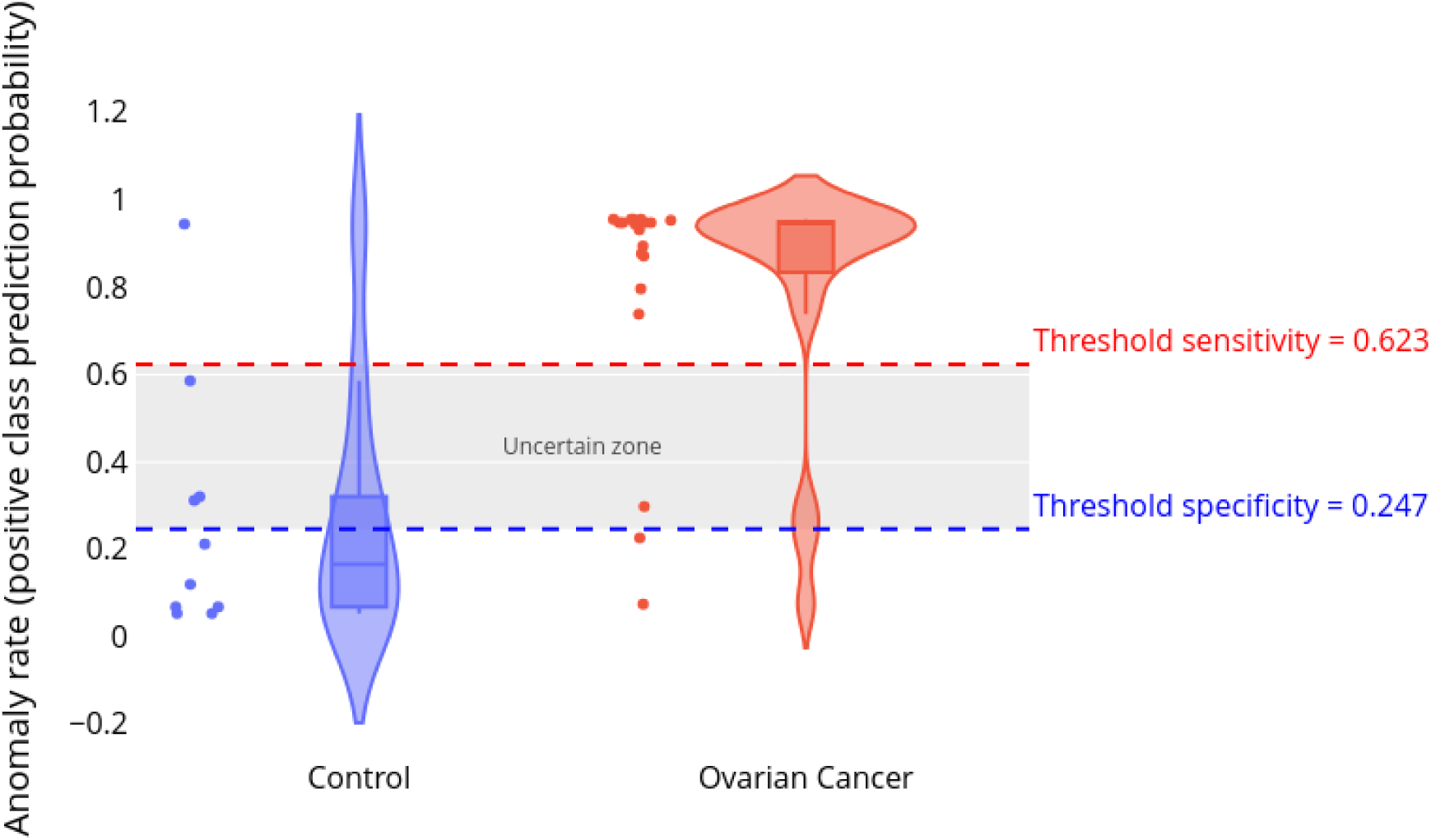
Performance of the combined-feature model on the test set.

Detailed metrics for each feature group, model group, and uncertainty zone configuration are summarized in Table 2, which provides a comprehensive overview of accuracy, sensitivity, specificity, AUC, and proportion of excluded samples.

**Table 2.**
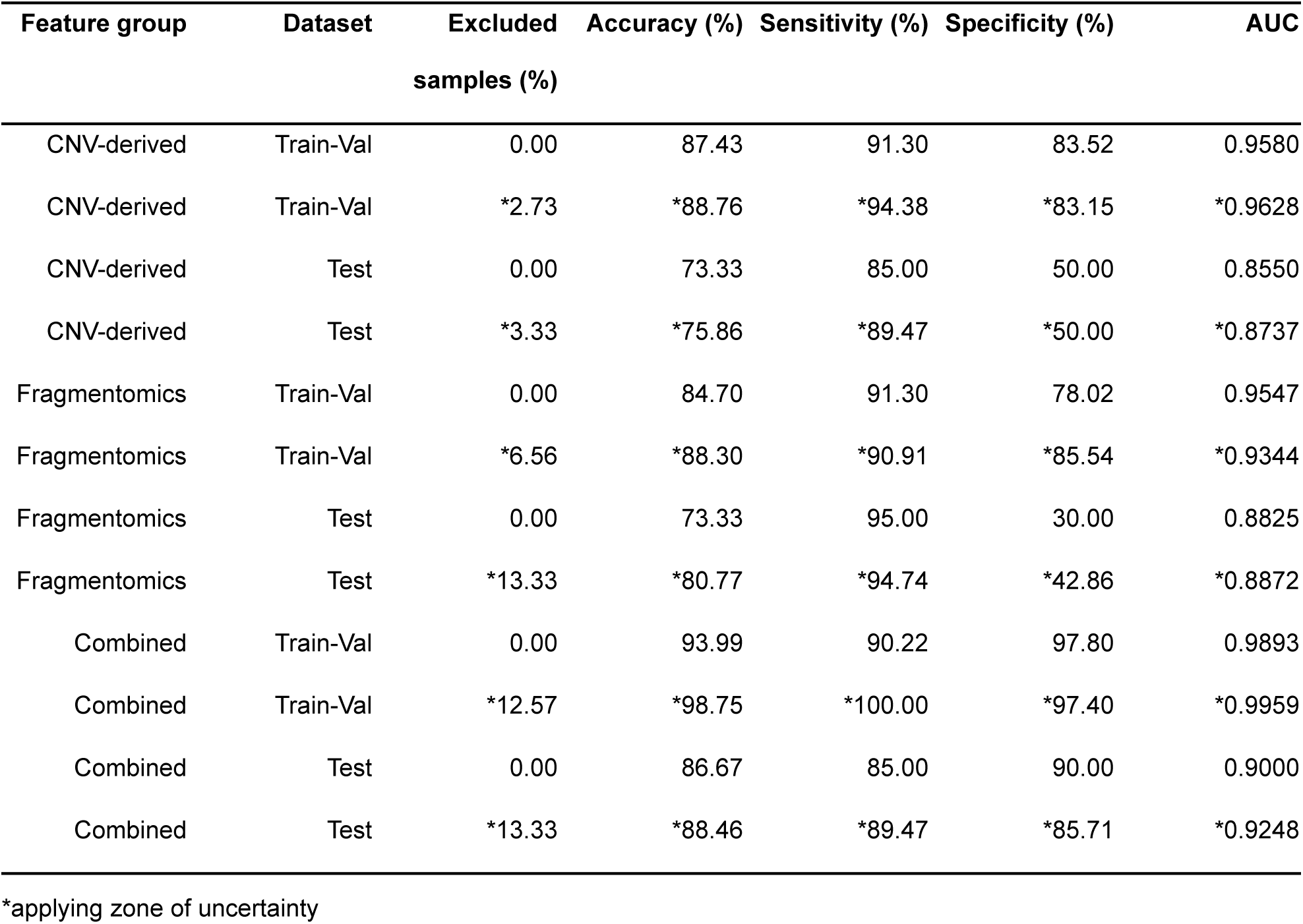
Summary of cfDNA ulcWGS-derived features used in the study.

| Feature group | Dataset | Excluded samples (%) | Accuracy (%) | Sensitivity (%) | Specificity (%) | AUC |
| --- | --- | --- | --- | --- | --- | --- |
| CNV-derived | Train-Val | 0.00 | 87.43 | 91.30 | 83.52 | 0.9580 |
| CNV-derived | Train-Val | *2.73 | *88.76 | *94.38 | *83.15 | *0.9628 |
| CNV-derived | Test | 0.00 | 73.33 | 85.00 | 50.00 | 0.8550 |
| CNV-derived | Test | *3.33 | *75.86 | *89.47 | *50.00 | *0.8737 |
| Fragmentomics | Train-Val | 0.00 | 84.70 | 91.30 | 78.02 | 0.9547 |
| Fragmentomics | Train-Val | *6.56 | *88.30 | *90.91 | *85.54 | *0.9344 |
| Fragmentomics | Test | 0.00 | 73.33 | 95.00 | 30.00 | 0.8825 |
| Fragmentomics | Test | *13.33 | *80.77 | *94.74 | *42.86 | *0.8872 |
| Combined | Train-Val | 0.00 | 93.99 | 90.22 | 97.80 | 0.9893 |
| Combined | Train-Val | *12.57 | *98.75 | *100.00 | *97.40 | *0.9959 |
| Combined | Test | 0.00 | 86.67 | 85.00 | 90.00 | 0.9000 |
| Combined | Test | *13.33 | *88.46 | *89.47 | *85.71 | *0.9248 |

\*applying zone of uncertainty

To enhance interpretability, SHAP analysis was performed on the combined model (Fig. 9). Feature attribution results demonstrated that both CNV-derived and fragmentomic metrics contributed substantially to predictions. High-impact CNV features were primarily associated with increased genomic instability patterns, whereas influential fragmentomic features reflected altered fragmentation length distributions and regional coverage variability. Importantly, the relative contribution of specific features varied across individual samples, supporting the presence of biological heterogeneity within OC cases. No single feature class dominated model output, indicating that classification performance arises from integrated, multidimensional signals rather than reliance on a single biomarker.

**Fig 9.**
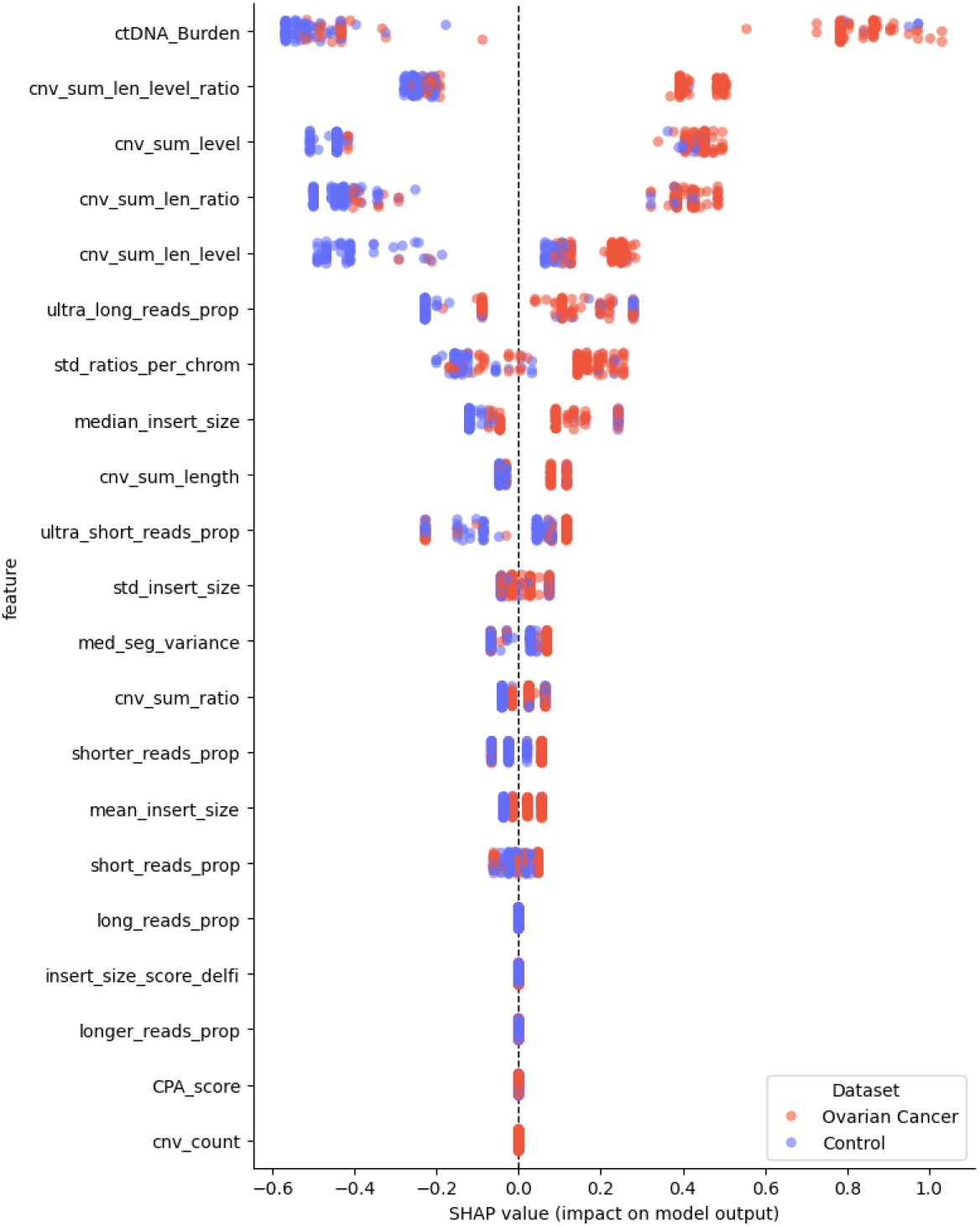
SHAP summary plot of the integrated model. The plot displays the contribution of features ranked by mean absolute SHAP value. Each point represents an individual sample, with its horizontal position indicating the SHAP value (impact on model output). Positive SHAP values correspond to a higher predicted probability of the oncologic class, whereas negative values indicate a shift toward the non-oncologic class. Points are color-coded according to the true class labels (e.g., Control vs Ovarian Cancer samples). The distribution of points reflects both the magnitude and direction of each feature’s contribution across the cohort, highlighting class-specific patterns, as well as the overall importance and variability of predictors in the integrated model.

Together, these results demonstrate that CNV and fragmentomic metrics encode complementary and partially interdependent information. Their integration yields a structured yet heterogeneous feature space in which disease-associated signals emerge through coordinated multidimensional patterns. Supervised modeling of the combined feature set translates these complex molecular relationships into improved diagnostic discrimination, supporting the value of integrated cfDNA profiling for OC detection.

### Evaluation of follow-up samples

The evaluation of follow-up samples using two best-performing model metrics demonstrated strong discriminative performance relative to the healthy control group (Fig. 10). For each patient, values were averaged across all available samples (separately for baseline and follow-up) to obtain a single representative score. The best-performing metric, ctDNA_Burden (Fig. 10A), achieved an AUC of 0.810 for pre-surgery samples, while follow-up samples reached a slightly lower AUC of 0.753, indicating that although the signal is partially reduced after surgical intervention, the metric retains substantial ability to distinguish oncological samples from healthy individuals. The second-best metric, cnv_sum_len_level_ratio (Fig. 10B), showed a comparable trend, with an AUC of 0.905 for pre-surgery samples and 0.843 for follow-up samples.

**Fig 10.**
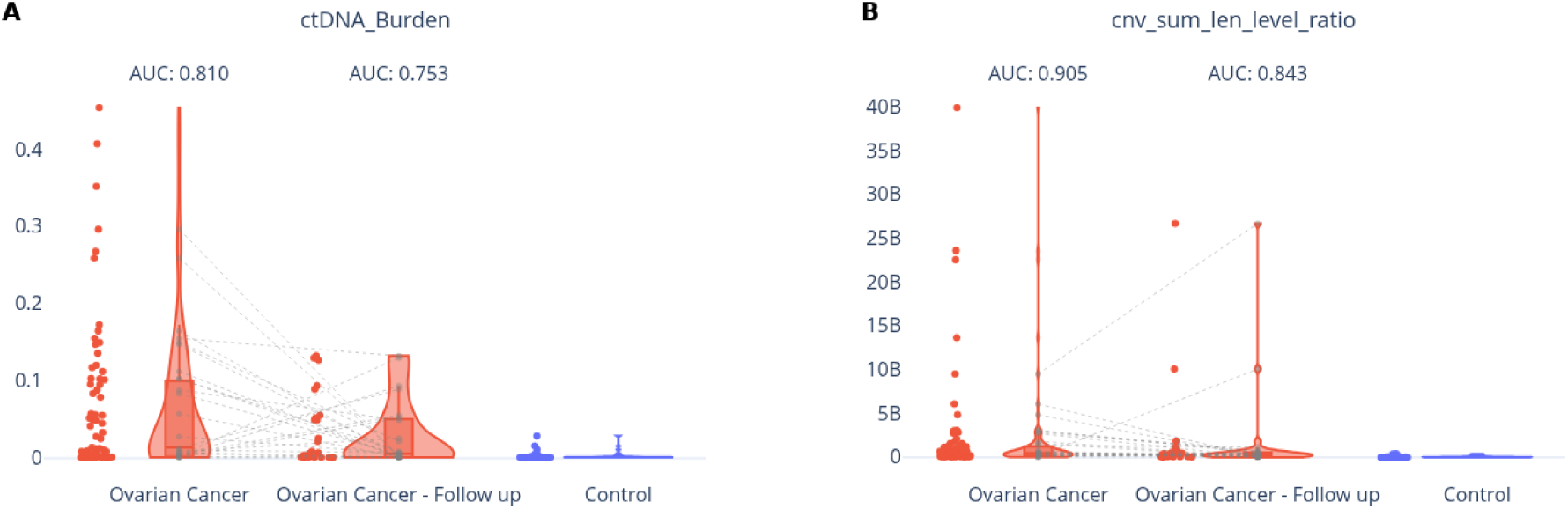
Comparison of model performance patient-averaged metrics in the oncological cohort between preoperative and postoperative samples. Panel A shows the best-performing metric -ctDNA_Burden, while Panel B presents the second-best model metric -cnv_sum_len_level_ratio. Dashed lines indicate paired connections between pre-surgery and follow-up (post-surgery) samples from the same individuals.

## Discussion

In this study, we developed and evaluated an integrated ulcWGS framework for OC detection that combines CNAs reflecting macroscopic genomic instability with cfDNA fragmentation patterns reflecting nucleosome-associated chromatin structure. The model captures complementary tumor-associated signals within an affordable framework. Interpretability analyses (Fig. 9) confirmed that classification is driven by coordinated contributions of both features types rather than a single dominant metric. Although CNV- and fragmentomics-derived features showed measurable group-level differences, substantial overlap at the sample level (Fig. 2 – 5; Table 2) indicates that disease-associated signals are distributed across multiple cfDNA layers and are more effectively captured when integrated. This observation is consistent with previous studies demonstrating that combining cfDNA-derived features improves classification performance in low-coverage WGS settings. For example, Liu et al. [52] showed that a model combining CNV, fragment size distribution, and nucleosome footprint features outperformed individual base models. Similarly, single-feature approaches such as CNV or fragment length alone showed lower performance (e.g., AUC ∼0.76 for CNV and ∼0.61 for fragment length) [53], while previous studies indicate that individual cfDNA biomarkers often have limited sensitivity or specificity at low tumor fractions, whereas feature integration improves robustness [54].

In our data, a similar pattern was observed. The fragmentomics-only model achieved high sensitivity but low specificity, whereas the CNV-only model showed moderate performance. The combined model provided a more balanced result, with improved specificity in the independent test set (Table 2). These findings suggest that individual cfDNA feature groups capture only part of the disease-associated signal, while their integration reduces false positives and improves overall discrimination. This is particularly relevant in OC, where ctDNA is often present at low fractions and tumors exhibit widespread chromosomal instability, which limits the performance of single biomarker approaches. Genome-wide approaches such as ulcWGS are therefore well suited for capturing these signals, whereas targeted assays may miss a substantial proportion of cases. Consistent with this, CNV-based cfDNA profiles derived from ulcWGS can distinguish OC patients from healthy individuals even at low sequencing depths [17, 55], while fragmentomic alterations provide complementary information reflecting tumor-associated chromatin changes [26].

At the same time, the benefit of feature integration is context-dependent. Helzer et al. [56] demonstrated that the performance of individual fragmentomic features depends on cancer type and assay design, and that combining all features does not always improve results. Thus selecting the optimal fragmentomics metrics can have a large effect on performance. Our findings therefore suggest that, in OC and under ultra-low coverage WGS conditions, combining CNV, ctDNA burden, and fragmentation-related metrics can provide a more robust signal than using a single feature group alone.

Because OC typically progresses without specific symptoms and lacks effective screening strategies [12], most cases are identified at advanced stages associated with significantly reduced survival. There is evidence that ctDNA levels correlate with tumor burden and clinical stage, showing markedly lower concentrations in early-stage disease and substantially higher levels in advanced tumors [57]. Consistently, a meta-analysis by Lu & Li (2021) [58] reported that elevated ctDNA is also associated with worse overall and progression-free survival, particularly in advanced FIGO stages. Our framework reliably distinguished OC patients spanning FIGO stages I – IV from healthy controls (Fig. 8; Table 2). Although the number of early-stage cases was limited (*n* = 14) (S1 Fig.), these findings suggest that the method remains informative even when ctDNA fractions are expected to be low. This likely reflects the integration of both macroscopic and microscopic cfDNA signals, as also supported by the combined feature space analysis, where integration of CNV and fragmentomic features resulted in improved but still partially overlapping separation of OC and control samples (Fig. 6), enabling detection of subtle tumor-associated deviations rather than relying solely on the high ctDNA abundance typically observed in advanced disease.

Therapeutic interventions themselves may also substantially influence cfDNA profiles. In advanced epithelial OC, an early decrease of ctDNA after a single cycle of neoadjuvant chemotherapy has been shown to be strongly associated with improved progression-free and overall survival. This rapid decline likely reflects an early cytotoxic or cytostatic effect leading to reduced tumor-derived DNA release into circulation [59]. Given the short half-life of cfDNA, such treatment-induced changes may occur within days and significantly alter the relative proportion of tumor-derived fragments within the total cfDNA pool. Beyond quantitative reductions, systemic therapy may also modify structural properties of cfDNA. In breast cancer, neoadjuvant chemotherapy has been reported to significantly shorten cfDNA telomere length, particularly in treatment-responsive patients [60], suggesting that therapy can reshape the molecular composition of cfDNA. However, chemotherapy-induced changes in total cfDNA do not necessarily translate into proportional alterations in tumor-specific genomic features, as systemic therapy may affect overall cfDNA dynamics and tumor-derived CNA profiles can remain relatively stable [61].

The observed decrease in performance on the independent test set compared to the train-validation cohort (Table 2; Fig. 8 – 9) may be attributed to several non-technical factors. While the integrated model achieved high performance in the training-validation cohort (AUC 0.989; sensitivity 90.22% and specificity 97.80%), its performance in the independent test set was more modest (AUC 0.900; sensitivity 85% and specificity 90%), indicating that model performance did not fully translate to the independent test set. First, biological heterogeneity of OC, including variability in tumor burden and molecular subtypes (S1 Fig.), may result in cfDNA patterns that differ from those represented during model optimization. Second, the potential presence of non-malignant pathologies in the control group may introduce confounding cfDNA alterations that reduce specificity. However, detailed clinical information regarding comorbid conditions in control individuals was not available in our cohort, and therefore their potential contribution to cfDNA variability could not be systematically assessed. This consideration is particularly relevant because a number of non-oncological disorders are known to influence cfDNA characteristics. Chronic inflammatory, autoimmune, and metabolic conditions, such as inflammatory bowel disease (IBD), diabetes mellitus, acute infections, ischemic events, obesity, and systemic stress, have been shown to alter both cfDNA concentration and fragmentation dynamics [62, 63]. For instance, patients with diabetes mellitus complicated by diabetic nephropathy exhibit elevated cfDNA levels, a reduction in long fragments (>250 bp), and an increased proportion of nucleosome-sized fragments (160 – 170 bp), accompanied by distinct end-motif alterations [64]. Similarly, individuals with IBD, even during clinical remission, demonstrate increased circulating nuclear and mitochondrial cfDNA compared with healthy controls, despite unchanged total cfDNA levels [65]. Such inflammation and tissue injury-related processes may generate cfDNA profiles that partially overlap with tumor-associated signatures, contributing to false-positive classifications and the lower specificity observed in fragmentomics-only (30.00%) and CNV-only (50.00%) models but less so in the combined model (90.00%) (Table 2). These findings highlight that cfDNA fragmentomic signals reflect broader biological processes beyond malignancy and underscore the importance of accounting for non-oncological variability when developing screening-oriented classifiers, as also reflected by the partially overlapping cluster structures observed in the integrated feature space (Fig. 7). Finally, given the limited size of the test cohort, performance estimates are sensitive to single-sample misclassification, where even one incorrectly classified sample may substantially affect percentage-based metrics.

The most informative features capture complementary aspects of cfDNA biology, including structural genomic instability and fragmentation irregularities. CNV-based metrics such as the number and cumulative length of CNV events reflect the extent of large-scale genomic disruption, a hallmark of tumor-derived cfDNA and a fundamental feature of genomic variability and disease-associated structural alterations [18]. On the other hand, fragmentation-focused attributes capture tumor-associated alterations in nucleosomal organization and nuclease activity. Together, these metrics form a coherent multidimensional signature that differentiates OC from healthy cfDNA profiles. Importantly, the relative contribution of specific features varied across individual samples (Fig. 9), supporting the presence of biological heterogeneity within OC cases. Part of this variability may also arise from the ultra-low sequencing depth, which introduces sampling-related variabilities in both CNV and fragmentomic features. The observed differences therefore likely reflect a combination of biological diversity and data-related variability (Fig. 2 – 7, 9).

From a screening perspective, the balance between sensitivity and specificity is critical. Screening tests inherently prioritize sensitivity, as the primary goal is to identify all individuals who may have the disease. In practical terms, we accept a certain level of false positivity, which may lead to additional follow-up examinations for a small number of individuals, while missing an oncological case, particularly in HGSOC, would undermine the fundamental purpose of early detection. The design and evaluation of our model thus emphasize high sensitivity to ensure that the vast majority of patients with cancer are captured by the screening pipeline.

In addition to therapy-induced changes, surgical intervention represents a distinct biological context influencing cfDNA dynamics. In our cohort, analysis of post-surgery follow-up samples (Fig. 10) demonstrated a partial shift of cfDNA profiles toward healthy-like patterns, consistent with a reduction of tumor-derived signal after tumor removal. Notably, a subset of follow-up samples was directly matched to corresponding baseline samples, enabling limited longitudinal comparisons, while additional samples were available only as standalone measurements. However, discriminative performance remained relatively high (AUC = 0.833 vs. 0.864 pre-surgery), indicating that tumor-associated features are not fully eliminated. This persistence may reflect residual disease, ongoing systemic effects, or delayed normalization of cfDNA fragmentation and CNV profiles. Interpretation of these results is further limited by the heterogeneity of follow-up sampling, including variable time intervals after surgery, differences in treatment regimens, and limited longitudinal depth per patient. Nevertheless, these findings suggest that cfDNA-based metrics capture dynamic, treatment-associated changes and may have potential utility beyond initial detection, particularly for disease monitoring and early identification of recurrence.

Therefore, our findings should be interpreted as a proof of concept that requires further validation in independent and clinically diverse cohorts. In conclusion, despite the limitations, the main contribution of this study lies in demonstrating that meaningful discrimination can be achieved using an ultra-low coverage (∼1×) cfDNA workflow without the need for deep sequencing or multi-omic assays, while operating at substantially lower sequencing depth than many reported approaches, often closer to 3 – 5× [66, 67] and targeted sequencing panels [56], this strategy represents an affordable and scalable alternative for cfDNA-based cancer detection. The improved balance between sensitivity and specificity further supports the concept that integrating complementary biological signals may be more beneficial than increasing sequencing depth alone, particularly in the context of future screening-oriented applications.

## Declarations

### Ethics approval and consent to participate

The present study was conducted in accordance with the ethical guidelines of all participating institutions and adhered to established international and national ethical standards. The study protocol was prepared by Clinomics Europe Ltd. and approved by the Ethics Committee of the National Center for Public Health and Pharmacy (Study approval number: 16119-8/2022/EÜIG; Ethics committee reference number: IV/1660-5/2022/EKU). All procedures were carried out in accordance with the Declaration of Helsinki and applicable regulatory requirements. To safeguard participant confidentiality, a double-blind design was implemented: data analysts responsible for generating and processing sequencing data were blinded to clinical information, while clinical personnel did not have access to analytical results. Clinical and pathological annotations were stored and managed using the Research Electronic Data Capture (REDCap) system [68, 69] hosted at the Comenius University in Bratislava, ensuring secure, access-controlled data governance.

### Consent for publication

Not applicable, as this manuscript does not contain any individual person’s identifiable data.

## Author information

### Author contributions

Z.H.: conceptualization, methodology, validation, visualization, data curation, software, formal analysis, writing -original draft preparation and revision. J.St.: conceptualization, visualization, writing -original draft preparation and revision, project administration. O.P.: conceptualization, visualization, writing -original draft preparation and revision. O.B.: clinical sample provision, project administration (ethical approval and regulatory compliance), review and editing. S.B.: investigation (cfDNA extraction, library preparation, sequencing). L.L.: investigation (cfDNA extraction, library preparation, sequencing). J.Sit.: methodology and software. T.Sl.: methodology and software. W.K.: methodology and software. A.M.: investigation (plasma processing and sample preparation), data curation. P.H.: methodology and software. Sz.M.: clinical sample collection, data curation. Á.E.: clinical sample collection, data curation. J.R.: clinical sample collection, data curation. T.Sed.: supervision. J.Radv.: conceptualization, supervision. J.B.: conceptualization, methodology, investigation, writing -original draft, review and editing. T.Sz.: supervision, funding acquisition, project administration. All authors read, revised and approved the final manuscript.

## Acknowledgments

We acknowledge the contributions of collaborating institutions involved in this study: the First Department of Obstetrics and Gynaecology, Semmelweis University (sample collection); Comenius University in Bratislava, hosted by the Comenius University Science Park (whole-genome sequencing analysis and data analysis); Clinomics Europe Ltd. (sample preparation and provision); and Geneton Ltd. (IT solutions and bioinformatics support).

## Data availability statement

Data generated and analyzed during this study are included in this published article and its supplementary information files. Due to privacy and ethical restrictions related to patient data, raw sequencing data are not publicly available but can be accessed from the corresponding author upon reasonable request and subject to institutional and ethical approval.

## List of abbreviations

AUC: Area under the receiver operating characteristic curve
cfDNA: cell-free DNA
ctDNA: Circulating Tumor DNA
CNA(s): copy number alteration(s)
CNV(s): copy number variation(s)
FIGO: international federation of gynecology and obstetrics
EOvC: endometrioid ovarian cancer
HGSOC: high-grade serous ovarian cancer
LGSOC: low-grade serous ovarian cancer
ML: machine learning
OC: ovarian cancer
PCA: principal component analysis
PCR: polymerase chain reaction
SHAP: shapley additive explanations
ulcWGS: ultra-low coverage whole genome sequencing
WGS: whole genome sequencing

---

**S1 Fig.**
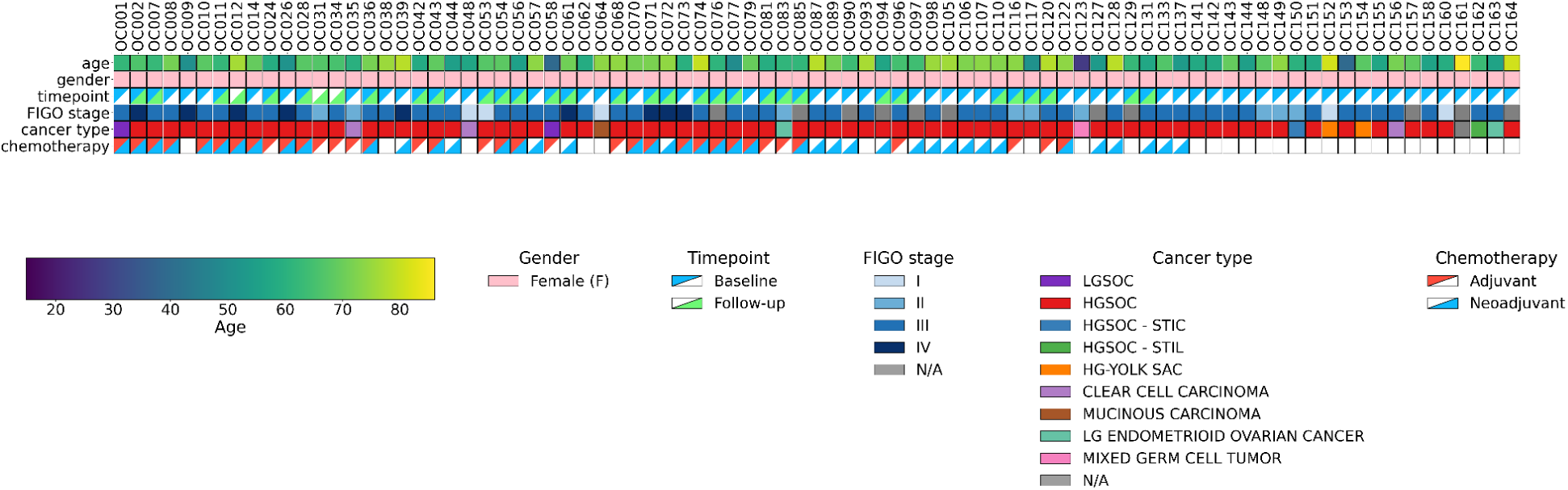
Cohort characterization. The figure summarizes the age distribution of participants at the time of their first blood sample collection and presents key clinicopathological characteristics of ovarian cancer patients, including FIGO stage, histological subtype, sample time point (baseline vs. follow-up), and treatment status with respect to neoadjuvant and adjuvant chemotherapy. Epithelial tumors comprise high-grade serous ovarian carcinoma (HGSOC), including cases associated with serous tubal intraepithelial carcinoma (HGSOC -STIC) and serous tubal intraepithelial lesion (HGSOC -STIL), as well as low-grade serous (LGSOC) ovarian cancer, clear cell carcinoma, mucinous carcinoma, and low-grade endometrioid ovarian cancer. Non-epithelial tumors include mixed germ cell tumors, high-grade yolk sac tumor (HG-YOLK SAC), and rare mixed tumors with combined HGSOC and yolk sac tumor components.

**S1 Table.**
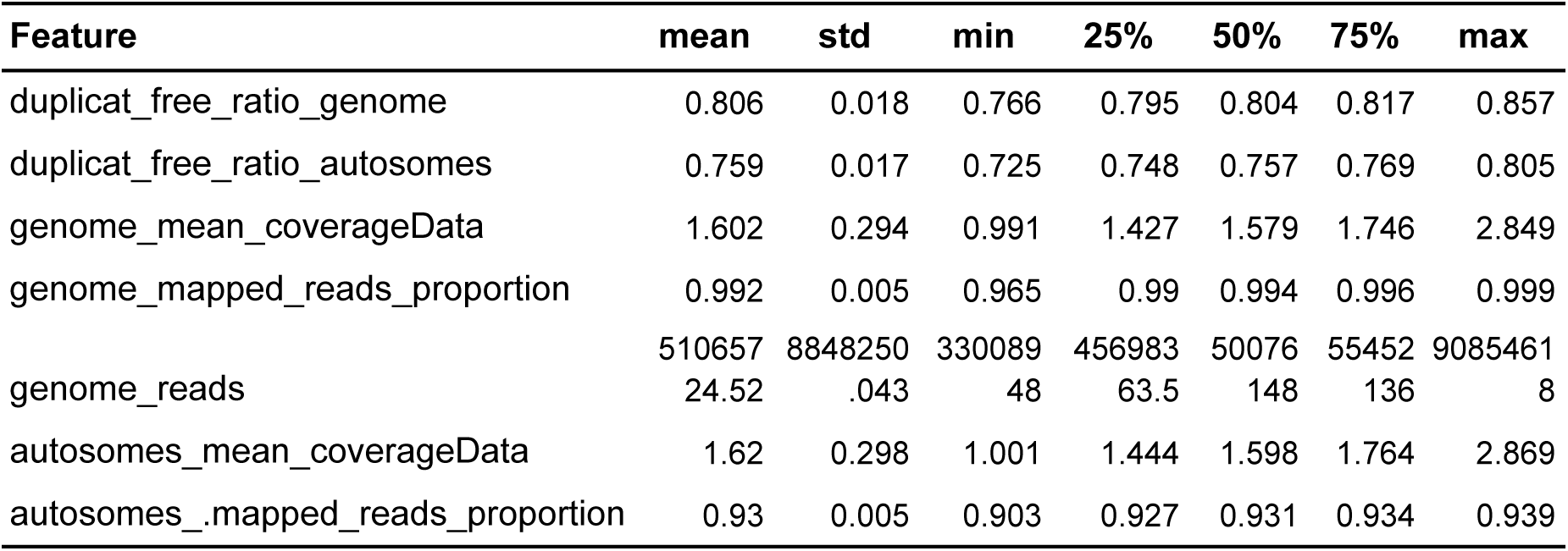
Samples coverage characterization. Descriptive statistics of selected coverage metrics on the whole genome and separately on autosomes.

| Feature | mean | std | min | 25% | 50% | 75% | max |
| --- | --- | --- | --- | --- | --- | --- | --- |
| duplicat_free_ratio_genome | 0.806 | 0.018 | 0.766 | 0.795 | 0.804 | 0.817 | 0.857 |
| duplicat_free_ratio_autosomes | 0.759 | 0.017 | 0.725 | 0.748 | 0.757 | 0.769 | 0.805 |
| genome_mean_coverageData | 1.602 | 0.294 | 0.991 | 1.427 | 1.579 | 1.746 | 2.849 |
| genome_mapped_reads_proportion | 0.992 | 0.005 | 0.965 | 0.99 | 0.994 | 0.996 | 0.999 |
| genome_reads | 510657<br>24.52 | 8848250<br>.043 | 330089<br>48 | 456983<br>63.5 | 50076<br>148 | 55452<br>136 | 9085461<br>8 |
| autosomes_mean_coverageData | 1.62 | 0.298 | 1.001 | 1.444 | 1.598 | 1.764 | 2.869 |
| autosomes_mapped_reads_proportion | 0.93 | 0.005 | 0.903 | 0.927 | 0.931 | 0.934 | 0.939 |

**S2 Fig.**
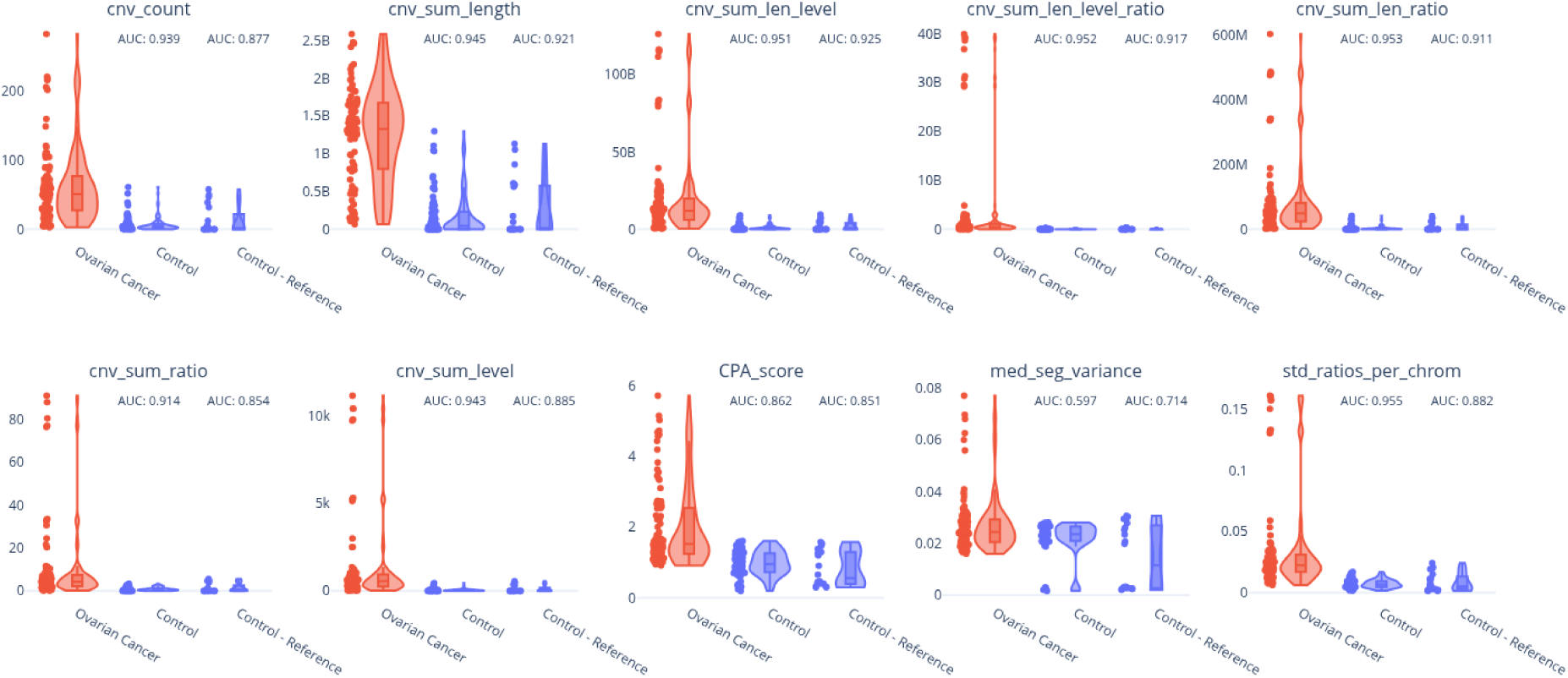
Violin plots of selected metrics comparing oncological samples (Ovarian Cancer) with two distinct healthy subgroups. Comparison of Control group (healthy controls excluding reference samples) and Control -Reference group (reference samples only). Each violin depicts the distribution of the respective metric within the indicated group, including overlaid boxplots and individual data points. The area under the receiver operating characteristic curve (AUC) is shown above each healthy subgroup and reflects the discriminative performance of the given metric for separating pre-surgery Ovarian Cancer samples from that specific subset of healthy controls.

